# What Factors are Important to the Success of Resubmitted Grant Applications in Health Research? A Retrospective Study of Over 20,000 Applications to the Canadian Institutes of Health Research

**DOI:** 10.1101/2024.05.29.24308137

**Authors:** J.G Wrightson, A. Lasinsky, R.R. Snell, M. Hogel, A. Mota, K.M. Khan, C.L. Ardern

**Author notes:** Corresponding author: Dr Clare Ardern City Centre 1 Building 13737 96 Avenue Surrey, BC V3V 0C6 Canada.

## Abstract

**Background:** In this retrospective study, we investigated the outcomes (funded/not funded) and factors related to the funding of resubmitted applications to the Canadian Institutes of Health Research (CIHR) Open Operating Grants Competition and Project Grant Competition between 2000 and 2022.

**Method and Findings:** The primary outcome was the proportion of resubmissions and new applications that were funded. Using a random forest model, we explored the importance of variables related to the success of resubmissions. A higher proportion of resubmissions (∼23%) were funded compared to new submissions (∼12%). The most important variables related to resubmission success were the rank (%) and score (/5) given to the preceding (initial) application and the number of the number of CIHR-funded grants where the PI was a named team member.

**Conclusion:** Resubmitting applications to the CIHR Project Grant Competition was beneficial, particularly for projects that were previously highly ranked and received high scores. These results may offer guidance for researchers who are deciding whether to resubmit rejected applications.

## Introduction

Few biomedical grant applications are successful [1]. When an application is rejected, applicants are typically encouraged to revise and resubmit their applications [2,3], an expensive and time-consuming task [4,5]. Given the low chances of success, researchers may question whether it is worth resubmitting the grant application [3]. Here, we describe the outcomes of resubmitted applications to the Canadian Institutes of Health Research (CIHR) Open Grants competitions.

In the U.S., resubmitted applications to the National Institute of Health (NIH) programs are more successful than new applications [2,6], though success rates remain modest [2]. Researchers require clear actionable information to help them decide whether to resubmit a rejected grant application [7], without which they may face multiple rejections from the same competition [8]. For NIH competitions, quantitative results (e.g. scores, ranking) from the peer review of the rejected application are related to whether someone decides to resubmit [9,10] and the success of the resubmission [11]. There is little data for other programs and countries, and it is not clear whether data from the NIH can be generalized to other funding systems [12]. Other factors such as applicants’ sex/gender, their previous funding success, and reviewer and review panel characteristics may also be related to whether a grant is funded [13,14]. Some of these factors may be related to how CIHR project grants are assessed [14]. However, to date, the factors related to the success of resubmissions to the CIHR project grant competition have not been explored.

Biomedical science and its benefits to society depend on successful applications for public research funding [15]. Substantial resources are wasted on repeated unsuccessful applications [4]. To help researchers decide whether to resubmit their rejected grant application, this exploratory study analyzed two decades of resubmission data from the CIHR Open Operating Grant and Project Grant competitions. The aims were to i) identify success rates of resubmitted CIHR operating and project grant applications and ii) explore the factors related to resubmission success.

## Method

This is a retrospective cohort study of observational data collected by CIHR between 2010 and 2022. The methods are reported in accordance with the relevant requirements of the Minimum Information about Clinical Artificial Intelligence Modeling (MI-CLAIM) checklist [16]. A completed checklist can be accessed online on the Open Science Framework (https://osf.io/pw45z/). The Behavioural Research Ethics Board at the University of British Columbia approved the study (H23-00719; March 31^st^, 2023).

### Context

The CIHR Open Grants competition–which currently accounts for approximately $750 million of CIHR’s $1.3 billion annual funding budget–is open to independent researchers at any career stage who seek funding to support their proposed fundamental or applied health-related research. The Open Grants competition comprises both the Open Operating Grants and the Project Grant competitions, data from which were extracted for 2010 to 2015 and 2017 to 2022, respectively. Each Peer Review Committee ranks the applications it considers in a competition, and an approach intended to account for varying ways that grant applications are scored by the approximately 60 different Peer Review Committees in each competition. The application rank, and not its score, dictates funding decisions.

### How CIHR handles resubmitted grant applications

An applicant may submit a previously unfunded application in a subsequent Open Grant competition round. There is currently no limit on the number of times an unsuccessful application can be resubmitted. Although applicants can provide a response to the previous application’s feedback, CIHR instructs peer review committee members to consider a resubmitted grant application as a new application (i.e. relative to all others in the current competition) and states that “…addressing previous reviews does NOT guarantee that the application will be better positioned to be funded…”. A resubmitted application can be reviewed by a different peer review committee than the previously unfunded application and committees do not have access to the previous version of the resubmitted application, though members are asked to read and evaluate the applicant’s response to the previous review. [17]

### Study design and dataset

Applications submitted to the Open Grant competition were surveyed to identify those that were new applications, unsuccessful, and followed by resubmission to the same program by the same Principal Investigator (PI). Data were extracted from 50138 applications to the CIHR Open Operating Grant and Project Grant competitions. Applications submitted to the 2016 Open Grant competitions were not included in this study because different reviewing and adjudication methodologies were used at that time. For the random forest model (see Data analysis, below), a randomly selected subset (20% of the data) was held back for model testing (“Test dataset”), and the remaining 80% of data were used for model training (“Training dataset”).

### Consent

Every researcher who submitted an application to the CIHR Project Grant Competition consented to the CIHR Policy on ‘Use of Personal Information’. CIHR maintained a record of all grant applications and the assessment records (including information from the Canadian Common Curriculum Vitae) of researchers who applied for funding. Our study included an objective of quality assurance and quality improvement of CIHR’s programs and thus fell under Article 2.5 of Tri-Council Policy Statement: Ethical Conduct for Research Involving Humans.

### Data analysis

We performed an exploratory analysis [18] to 1): Describe the proportion of new and resubmitted applications that were funded and 2) Identify the importance of variables related to the outcome (funded/not funded) of resubmissions using a random forest model. The primary outcome was the proportion of successful resubmitted and new grant applications (%).

### Data wrangling

The PI’s sex was hypothesized to be a significant explanatory variable [13]. Applications where the PI had not self-identified their sex (n = 37) were excluded. Applications were identified as resubmissions if they included a response to a previous review and could be paired with a preceding unfunded new application submitted by the same PI. The unit of analysis was application pairs. Resubmissions that could not be paired with a previously unfunded application (n = 9310) were excluded from analyses.

### Variable importance

We used a 10-fold cross-validated random forest model to identify the importance of candidate variables related to the outcome of resubmissions. Random forest is an ensemble learning model that combines multiple decision trees, makes few assumptions about distributions and inter-relationships between variables and can describe the relative importance of explanatory variables [19,20]. Plots showing the results of hyperparameter tuning are shown in the Supplementary Material, Figure S4. The parameter values selected maximized model sensitivity (true positive classification) and specificity (true negative classification) (mtry = 3, nodesize = 1, ntrees = 1000).

The relative importance of each explanatory variable was calculated using the mean decrease of Gini impurity (a method for assessing how effectively the explanatory variables split the data based on the primary outcome) aggregated across the folds. The primary aim of the analysis was explanation not prediction [21]. For completeness, the prediction accuracy of the model was established using the unseen (Test) dataset.

### Explanatory variables

The secondary outcome was the Gini importance of each of the candidate explanatory variables related to resubmission success. Candidate explanatory variables were selected from the factors related to grant resubmissions identified in our recent scoping review [12], other work highlighting possible biases in grant peer review selection [13,14], and from factors related to resubmissions in other funding competitions [9]). Two variable categories were identified: i) characteristics of the applicant (the named Principal Investigator (PI)) and ii) factors related to peer review. Variables included in the final model were: self-identified applicant sex (male, female), previous total CIHR funding awarded in CAD$M to all projects where the PI was a named team member (‘PI $ Funding’), the number of CIHR-funded projects that contributed to ‘PI $ Funding’ at the time of application (‘PI # grants’), whether the applicant had chosen

English or French as the application language, whether the resubmitted application was reviewed by the same peer review committee (true, false), whether the resubmitted application was reviewed by one or more of the same peer reviewers within the peer review committee (true, false), and the score (‘Previous Score, /5’) and ranking (‘Previous % Rank’) of the preceding application.

## Results

### Proportion of successful applications

The dataset included 40791 applications, 26142 of which were new applications and 14649 of which were resubmissions. During the study period, 3034 (∼12%) new applications and 3415 (∼23%) resubmissions were funded. Figure 1 shows density plots and median values for the scores and rankings for both new and resubmitted applications.

**Figure 1.**
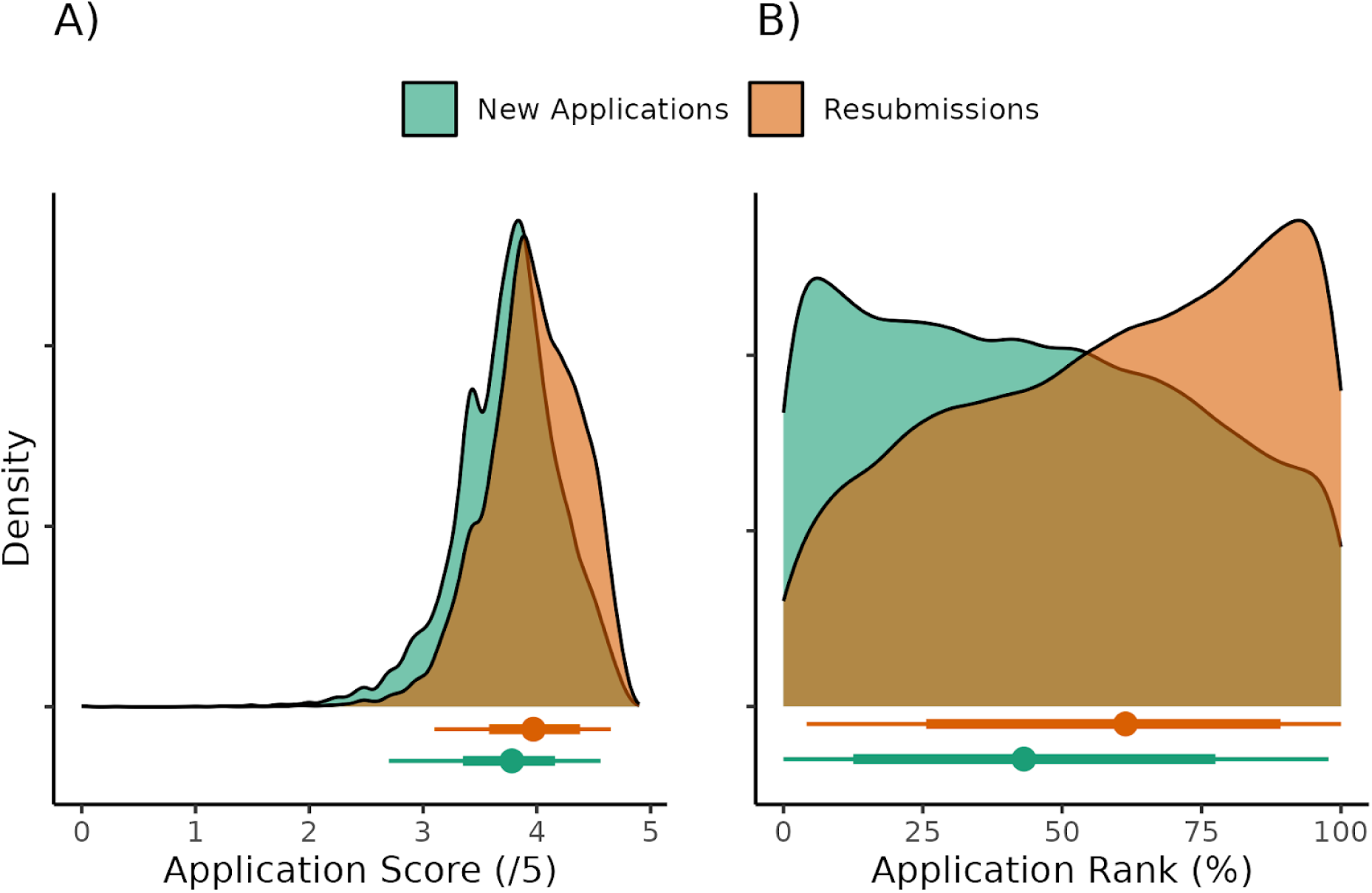
Density plots for the scores (panel A) and rank (panel B) for new and resubmitted applications. Circles and intervals represent the median ± the 66% and 95% quantiles

### Variables related to resubmission success

Table 1 shows descriptive statistics for the explanatory variables related to the Resubmission outcome (funded vs. unfunded) in both the Training and Test datasets. Relationships between candidate variables are shown in the supplementary material, Figures S1-S3. See ‘Explanatory variables’ above for a description of the variables entered into the model.

**Table 1.**
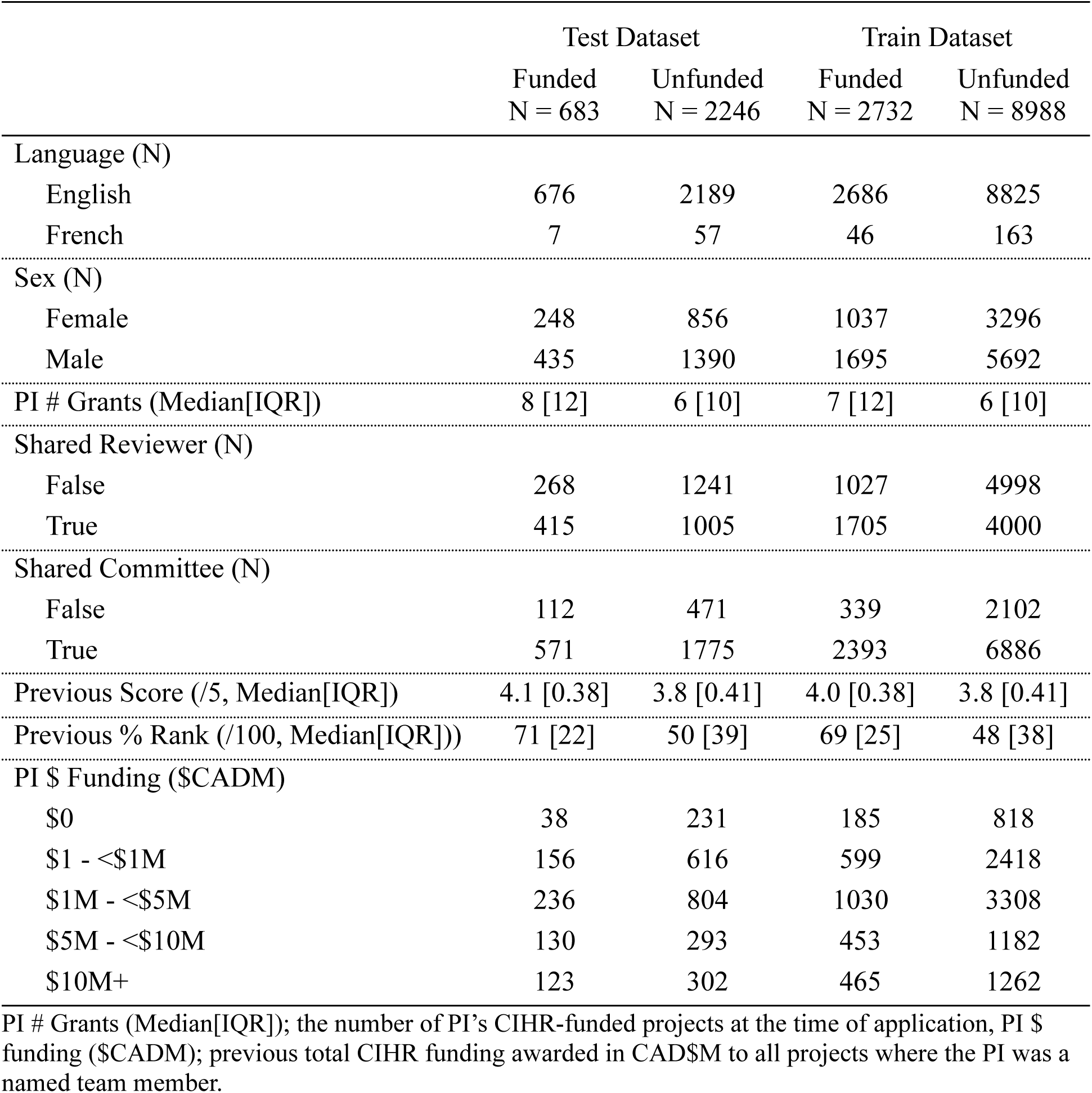
Explanatory variables related to resubmission outcome (funded vs. unfunded)

|  | Test Dataset |  | Train Dataset |  |
| --- | --- | --- | --- | --- |
|  | Funded<br>N = 683 | Unfunded<br>N = 2246 | Funded<br>N = 2732 | Unfunded<br>N = 8988 |
| Language (N) |  |  |  |  |
| English | 676 | 2189 | 2686 | 8825 |
| French | 7 | 57 | 46 | 163 |
| Sex (N) |  |  |  |  |
| Female | 248 | 856 | 1037 | 3296 |
| Male | 435 | 1390 | 1695 | 5692 |
| PI # Grants (Median[IQR]) | 8 [12] | 6 [10] | 7 [12] | 6 [10] |
| Shared Reviewer (N) |  |  |  |  |
| False | 268 | 1241 | 1027 | 4998 |
| True | 415 | 1005 | 1705 | 4000 |
| Shared Committee (N) |  |  |  |  |
| False | 112 | 471 | 339 | 2102 |
| True | 571 | 1775 | 2393 | 6886 |
| Previous Score (/5, Median[IQR]) | 4.1 [0.38] | 3.8 [0.41] | 4.0 [0.38] | 3.8 [0.41] |
| Previous % Rank (/100, Median[IQR]) | 71 [22] | 50 [39] | 69 [25] | 48 [38] |
| PI \$ Funding (\$CADM) | | | | |
| \$0 | 38 | 231 | 185 | 818 |
| \$1 - <\$1M | 156 | 616 | 599 | 2418 |
| \$1M - <\$5M | 236 | 804 | 1030 | 3308 |
| \$5M - <\$10M | 130 | 293 | 453 | 1182 |
| \$10M+ | 123 | 302 | 465 | 1262 |

The Gini variable importance data (from the training dataset) is shown in Figure 2. The percent rank assigned during peer review to the previous submission was the most important explanatory variable, followed by the previously assigned score (Figure 3). Application language (English, French) was the least important feature of the model.

**Figure 2.**
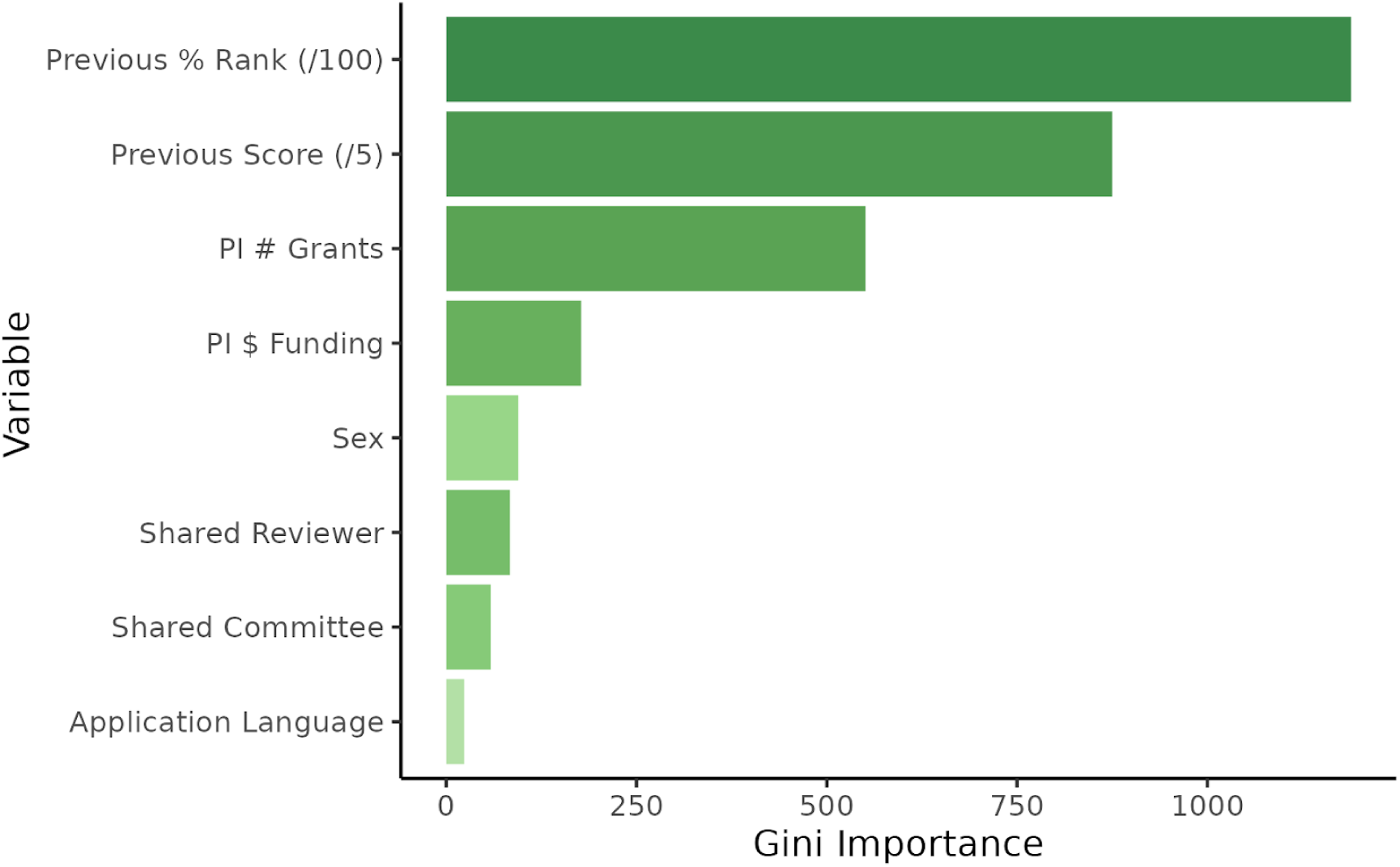
The candidate features entered into the random forest model, ranked by the mean decrease in Gini impurity (“Gini importance”). PI # Grants (Median[IQR]); the number of PI’s CIHR-funded projects at the time of application, PI $ funding ($CADM); previous total CIHR funding awarded in CAD$M to all projects where the PI was a named team member

**Figure 3.**
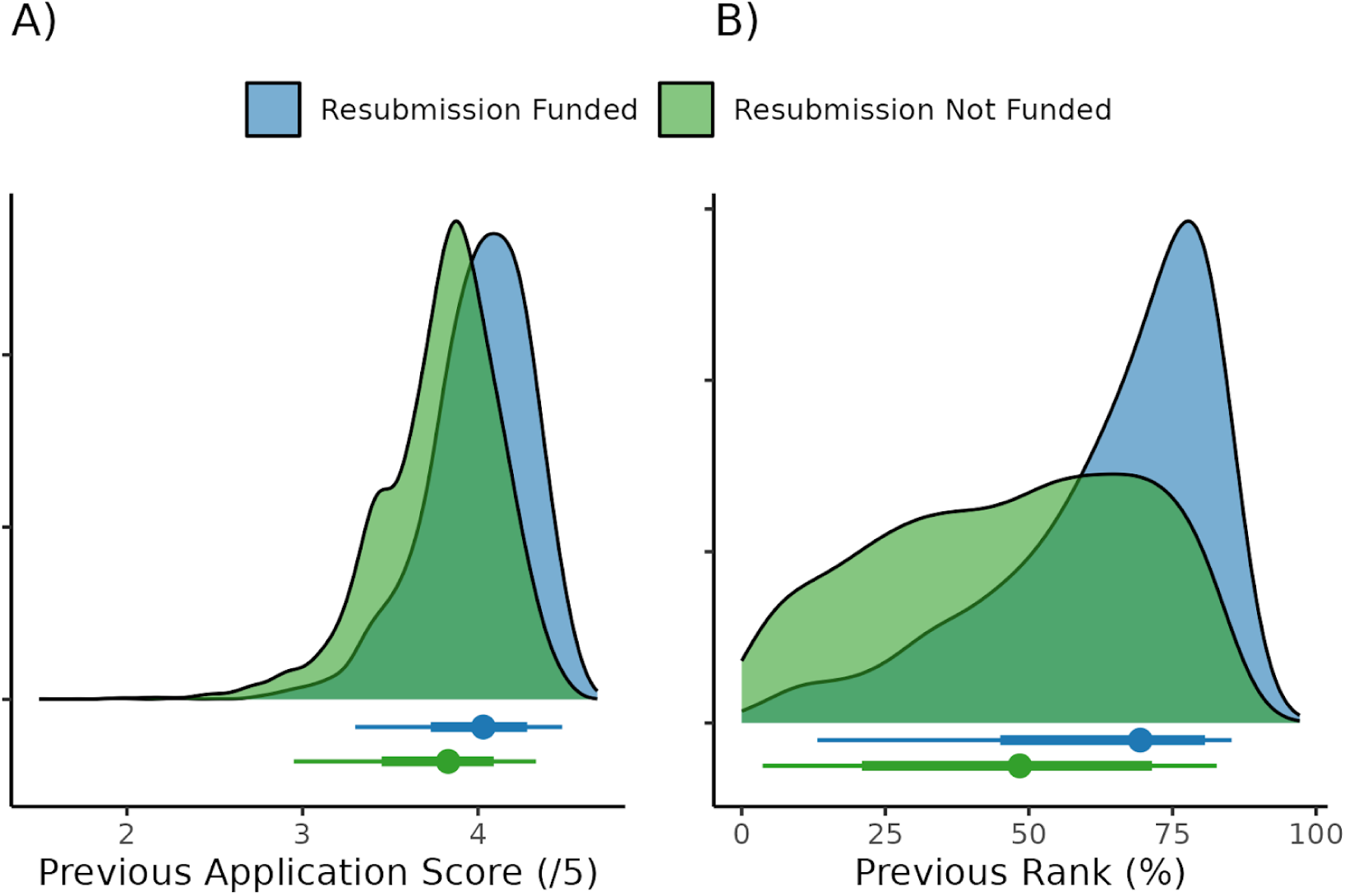
Density plots for the previous application’s score (Panel A) and rank (Panel B) for unfunded and funded resubmitted applications (combined for Test and Training datasets). Circles and intervals represent the median ± the 66% and 95% quantiles

### Prediction performance

The confusion matrix for the performance of the model in the Test dataset is shown in the Supplementary Material, Figure S5. The model classification accuracy was 0.76. As would be expected with such large class imbalances, the model was more accurate at predicting which resubmitted applications would be unsuccessful (specificity = 0.92) compared to which would be successful (sensitivity = 0.22).

## Discussion

Our results add to the literature on biomedical grant funding and peer review in two ways. First, we show that resubmitted applications to the Canadian Institutes of Health Research Project Grant Competition were funded more often and ranked and scored higher than new applications. Second, we show that the peer review ranking of the previous application was the most important variable related to whether a resubmitted application was funded. Applicant sex, application language and peer review continuity were the least important. Applicants who are considering resubmitting an unsuccessful application should feel encouraged by improved peer review outcomes for resubmissions, and may wish to consider the ranking of their previous application when deciding whether to resubmit a grant application to the CIHR Project Grant competition.

Matching data from the U.S. NIH [2,6], here we found a greater proportion of resubmitted applications were funded than new applications, and on average, resubmissions received a higher rank and score. We suggest three potential drivers of improved outcomes: First, it is possible that those who chose to resubmit were those who could adequately respond to the peer review feedback. Second, because CIHR treats resubmissions as new applications and instructs reviewers to compare them only to their present cohort, those who resubmit may be those whose application was more likely to be funded anyway (i.e. the application may be good but not quite good enough compared to the previous cohort it was initially judged against). Finally, a proportion of applicants whose applications received unfavourable reviews may choose not to resubmit, increasing the proportion of resubmissions which are likely to be funded.

Unlike journal peer review, providing applicant feedback is a lower priority of grant peer review committees [1,22]. However, applicants are recommended to, and do, use feedback to help with grant resubmission [7,10,23]. High-quality feedback helps researchers decide whether to submit their application [7]; low-quality feedback can confuse applicants [24] and may lead to multiple unsuccessful resubmissions [8]. CIHR has clear guidance for reviewers to promote high-quality reviews [25], which may have contributed to the improved success of resubmissions seen here. Unlike journal peer review [26], there has been little scientific evaluation of the impact of grant peer review feedback quality (but see also Derrick et al. 2023 [7]) perhaps due to the continuing opacity of many grant peer review systems, despite increasing calls for transparency [27–29]. Given the substantial time, effort and financial costs to applicants and society of grant applications, revisions and resubmissions [5,8,30], more research is warranted on how reviewer feedback impacts the volume, quality and success of subsequent grant applications.

Here we show that the most important factor related to the outcome of a resubmitted CIHR Open Grant was the rank given to the previous application. While many applicants may have *assumed* that the result of the previous peer review was related to the binary outcome of a resubmitted application, this is direct evidence of the relationship. The importance of rank compared to score may surprise some applicants, especially given the relevance of previous scores for resubmissions in other funding systems [9–11]. For some applicants, knowledge of the relationship between the previous applications’ rank and resubmission success may help them make an informed decision about whether to resubmit a grant application. We found that self-reported applicant sex was not an important factor related to resubmission outcome, mirroring the results of recent re-examinations of gender/sex bias in grant peer review [31,32].

The number of CIHR-funded projects the PI had been awarded was the third most important factor, after previous application rank and score. This result could be interpreted in at least two ways. The first is that grant writing is a skill [33], and one might assume that researchers may become more skilled in responding to reviewer comments with experience, especially with successful grants (Guyer et al., 2021). An alternative interpretation is that the result reflects the ‘Matthew effect’ in grant funding wherein previous success begets future success. A small group of previously successful researchers, rewarded more often on that basis, would threaten the assumed meritocracy of research funding systems [35]. Our observational data do not allow us to disentangle these explanations, though we speculate that both may be true. Recent innovations in grant peer review including ‘funding lotteries’ [36] and double-blind peer review processes [37] have been designed to reduce inherent bias in grant selection and peer review. Future research should examine whether the relationship between past success and the outcome of resubmissions still exists in applications to competitions that use these mechanisms designed to reduce bias.

This is an exploratory cross-sectional study, which precludes causal inferences about the relationship between the applicant and peer review characteristics, and grant resubmission success. We echo previous calls for further examination of grant peer review systems, including randomized controlled trials, to examine the causal factors that influence funding success [29,38,39]. We were unable to study the influence of many oft-reported biases in grant systems. For example, racial disparity in grant peer review and awards is well documented. However, because these data were not routinely collected by CIHR during the timeframe under consideration, we were unable to include self-identified race or ethnicity as factors in this analysis.

## Conclusion

Resubmitted applications to the CIHR Project Grant competition were, on average, funded more often and ranked higher than new submissions. The most important factor related to whether a resubmission was funded was the percent rank assigned to the previous unfunded application. Resubmission may be worthwhile, as long as the initial application was well reviewed and applicants can adequately respond to reviewer feedback. These data help increase the transparency of grant peer review and strengthen recent calls for increased scientific analysis of scientific funding systems [38–41].

## Code and data availability

Upon publication, the notebook containing the analysis code will be available on the Open Science Framework (https://osf.io/pw45z/). The data used in this analysis are held by CIHR and are not publicly available due to privacy and legal restrictions. Researchers wishing to obtain access to these data need to contact the Vice-President of Research Programs-Operations at CIHR to obtain approval to access de-identified data on operating grant funding program applications submitted between 2010 and 2022. Data for unfunded applications cannot be shared.

## Supplementary Material

**Figure S1.**
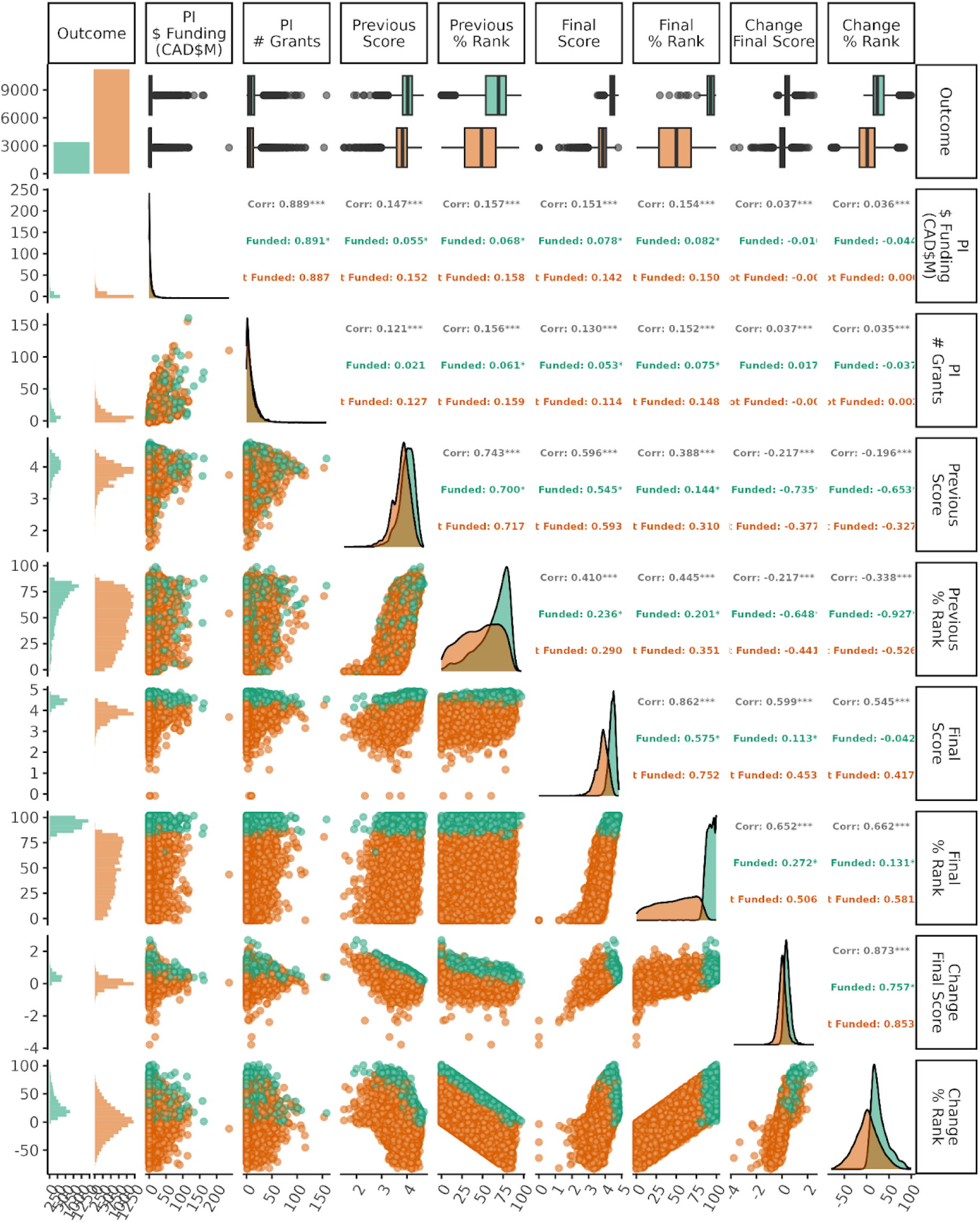
Relationships between continuous explanatory variables. The derivative variables 07 ‘Change in final score’ and ‘Change in % Rank’ are included for information only, they were 08 not candidate variables for the random forest analysis

**Figure S2.**
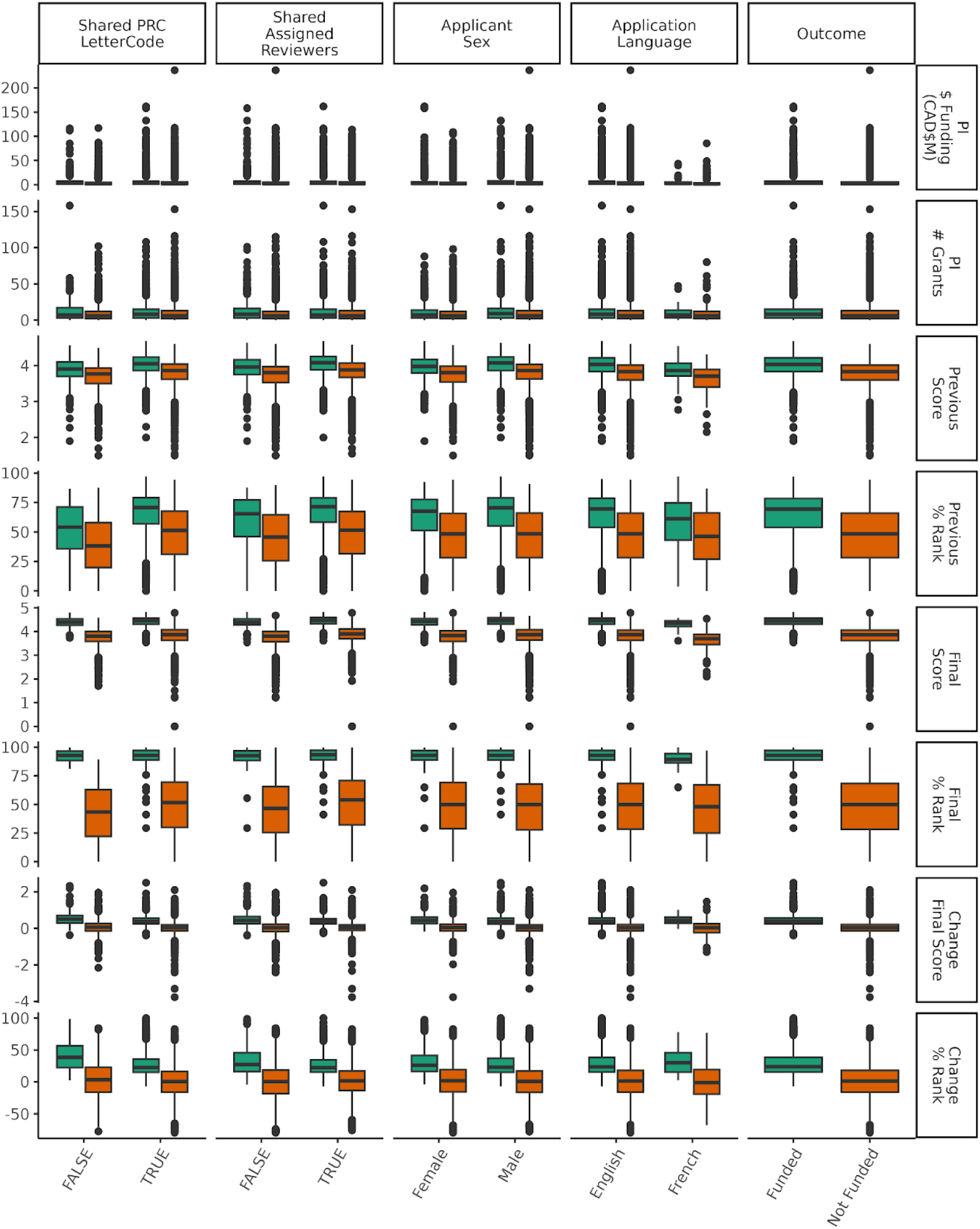
Relationships between continuous and categorical explanatory variables. The derivative variables ‘Change in final score’ and ‘Change in % Rank’ are included for information only, they were not candidate variables for the random forest analysis.

**Figure S3.**
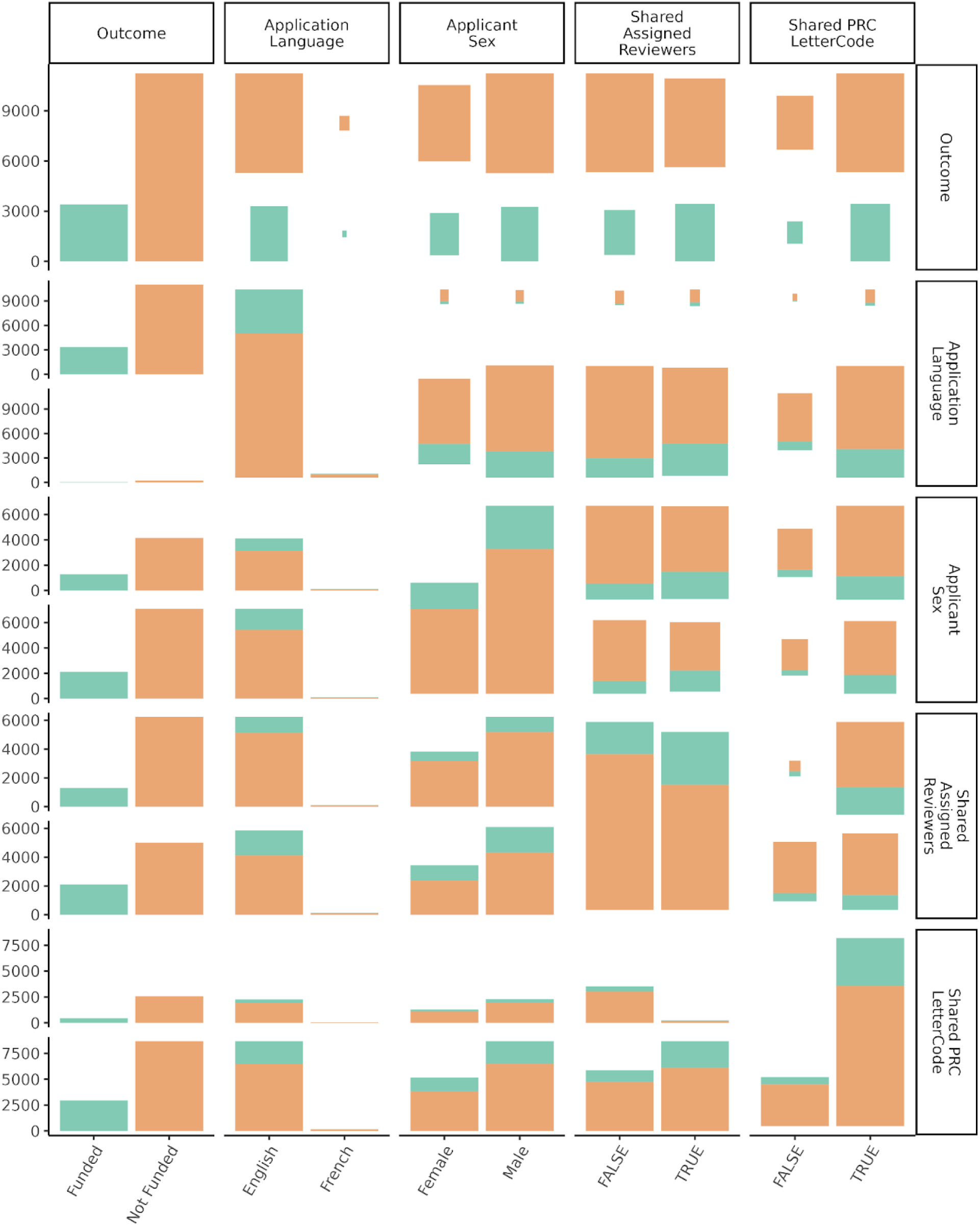
Relationships between categorical explanatory variables.

**Figure S4.**
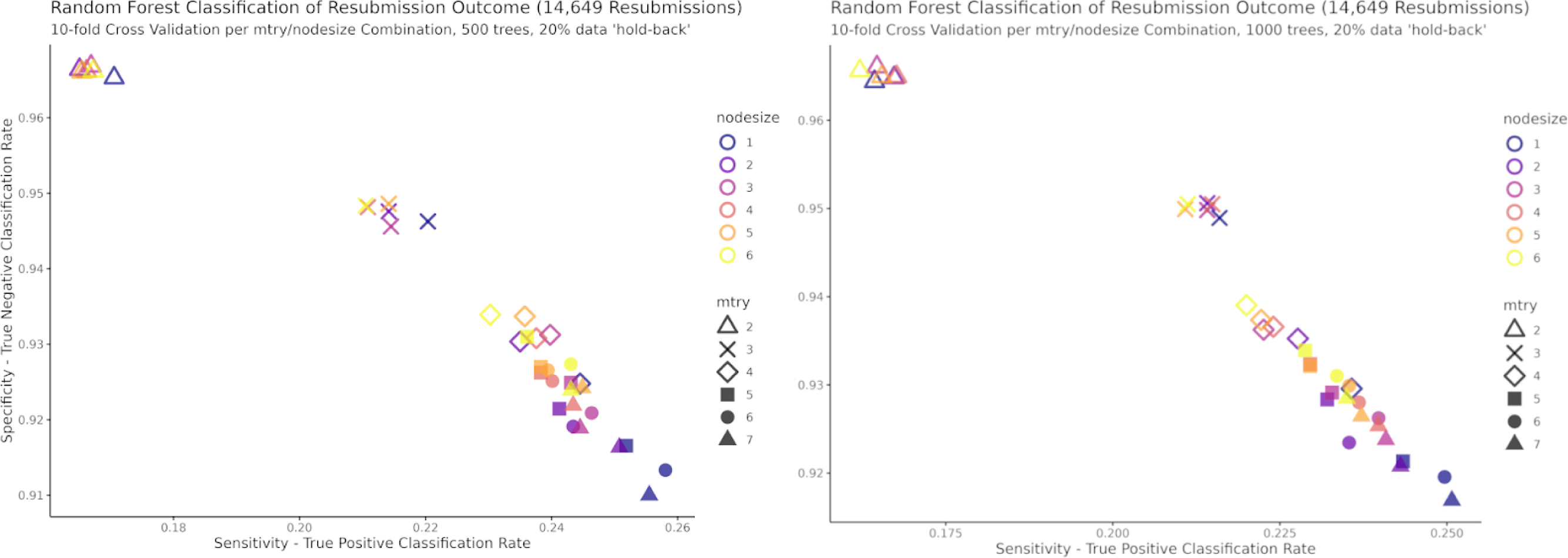
Effect of node size and *mtry* tuning on model sensitivity and precision for a model with 500 trees (left panel) and 1000 trees (right panel

**Figure S5.**
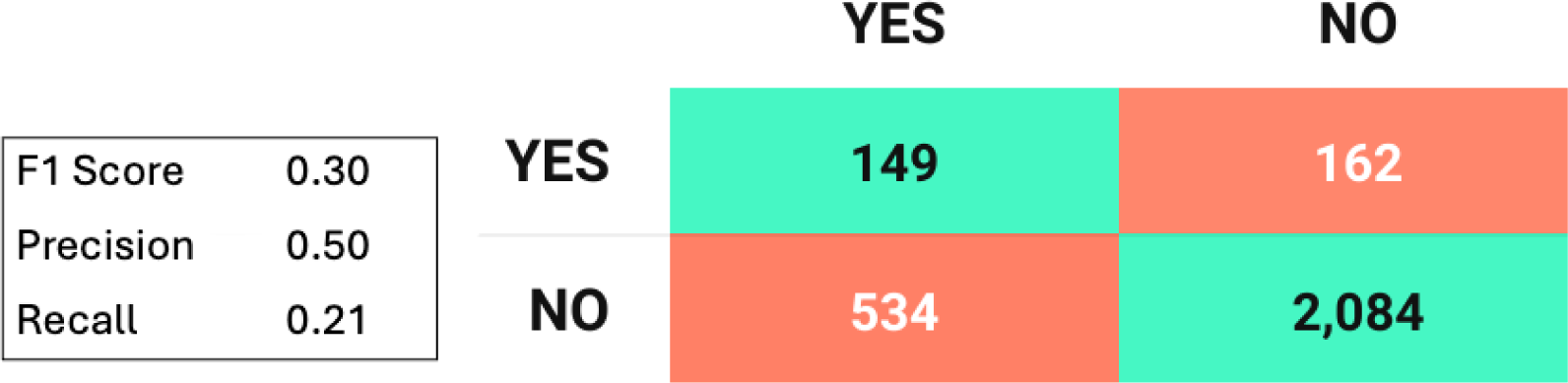
Confusion matrix for the random forest model performance in the Test dataset

## References

1. Guthrie S, Ghiga I, Wooding S. What do we know about grant peer review in the health sciences?: An updated review of the literature and six case studies. RAND Corporation; 2018 Jun. Available: https://www.rand.org/pubs/research_reports/RR1822.html

2. Lauer M. Are You On the Fence About Whether to Resubmit? 28 Oct 2016 [cited 30 Dec 2023]. Available: https://nexus.od.nih.gov/all/2016/10/28/are-you-on-the-fence-about-whether-to-resubmit/

3. Crow JM. What to do when your grant is rejected. Nature. 2020;578: 477–479. doi:10.1038/d41586-020-00455-0

4. Gross K, Bergstrom CT. Contest models highlight inherent inefficiencies of scientific funding competitions. PLOS Biol. 2019;17: e3000065. doi:10.1371/journal.pbio.3000065

5. Herbert DL, Barnett AG, Clarke P, Graves N. On the time spent preparing grant proposals: an observational study of Australian researchers. BMJ Open. 2013;3: e002800. doi:10.1136/bmjopen-2013-002800

6. Nakamura RK, Mann LS, Lindner MD, Braithwaite J, Chen M-C, Vancea A, et al. An experimental test of the effects of redacting grant applicant identifiers on peer review outcomes. Zaidi M, Isales C, editors. eLife. 2021;10: e71368. doi:10.7554/eLife.71368

7. Derrick GE, Zimmermann A, Greaves H, Best J, Klavans R. Targeted, actionable and fair: Reviewer reports as feedback and its effect on ECR career choices. Res Eval. 2023; rvad034. doi:10.1093/reseval/rvad034

8. von Hippel T, von Hippel C. To Apply or Not to Apply: A Survey Analysis of Grant Writing Costs and Benefits. PLOS ONE. 2015;10: e0118494. doi:10.1371/journal.pone.0118494

9. Boyington JEA, Antman MD, Patel KC, Lauer MS. Toward Independence: Resubmission Rate of Unfunded National Heart, Lung, and Blood Institute R01 Research Grant Applications Among Early Stage Investigators. Acad Med J Assoc Am Med Coll. 2016;91: 556–562. doi:10.1097/ACM.0000000000001025

10. Hunter CJ, Leiva T, Dudeja V. The unfunded grant, now what? Advice, approach, and strategy. Surgery. 2024;175: 317–322. doi:10.1016/j.surg.2023.09.057

11. Lauer M. Resubmissions Revisited: Funded Resubmission Applications and Their Initial Peer Review Scores. In: NIH Extramural Nexus [Internet]. 17 Feb 2017 [cited 4 Jan 2024]. Available: https://nexus.od.nih.gov/all/2017/02/17/resubmissions-revisited-funded-resubmission-applications-and-their-initial-peer-review-scores/

12. Lasinsky A, Wrightson J, Khan H, Moher P, Kitchin V, Khan KM, et al. If at First You Don’t Succeed: Biomedical Research Grant Resubmission Rates, and Factors Related to Success—A Scoping Review. Rochester, NY; 2024. doi:10.2139/ssrn.4803560

13. Guthrie S, Ghiga I, Wooding S. What do we know about grant peer review in the health sciences? F1000Research. 2018;6: 1335. doi:10.12688/f1000research.11917.2

14. Tamblyn R, Girard N, Qian CJ, Hanley J. Assessment of potential bias in research grant peer review in Canada. CMAJ. 2018;190: E489–E499. doi:10.1503/cmaj.170901

15. Yin Y, Dong Y, Wang K, Wang D, Jones BF. Public use and public funding of science. Nat Hum Behav. 2022;6: 1344–1350. doi:10.1038/s41562-022-01397-5

16. Norgeot B, Quer G, Beaulieu-Jones BK, Torkamani A, Dias R, Gianfrancesco M, et al. Minimum information about clinical artificial intelligence modeling: the MI-CLAIM checklist. Nat Med. 2020;26: 1320–1324. doi:10.1038/s41591-020-1041-y

17. Canadian Institutes of Health Research. Project Grant Program: Application Process. 30 Aug 2016 [cited 12 Feb 2024]. Available: https://cihr-irsc.gc.ca/e/49806.html#a2.2

18. Tukey JW. We Need Both Exploratory and Confirmatory. Am Stat. 1980;34: 23–25. doi:10.2307/2682991

19. Breiman L. Random Forests. Mach Learn. 2001;45: 5–32. doi:10.1023/A:1010933404324

20. Jones ZM, Linder FJ. edarf: Exploratory Data Analysis using Random Forests. J Open Source Softw. 2016;1: 92. doi:10.21105/joss.00092

21. Shmueli G. To Explain or to Predict? Stat Sci. 2010;25. doi:10.1214/10-STS330

22. Gluckman P, Ferguson M, Glover A, Grant J, Groves T, Lauer M, et al. International Peer Review Expert Panel report: A report to the Governing Council of the Canadian Institutes of Health Research. purpose; 2017 May. Available: https://cihr-irsc.gc.ca/e/50248.html

23. National Institute of Allergy and Infectious Diseases. Revise and Resubmit an Application. 16 Nov 2023 [cited 5 Jan 2024]. Available: https://www.niaid.nih.gov/grants-contracts/revise-resubmit-application

24. Gallo SA, Schmaling KB, Thompson LA, Glisson SR. Grant Review Feedback: Appropriateness and Usefulness. Sci Eng Ethics. 2021;27: 18. doi:10.1007/s11948-021-00295-9

25. Canadian Institutes of Health Research. Review Quality. 12 Jan 2018 [cited 5 Jan 2024]. Available: https://cihr-irsc.gc.ca/e/50787.html

26. Superchi C, González JA, Solà I, Cobo E, Hren D, Boutron I. Tools used to assess the quality of peer review reports: a methodological systematic review. BMC Med Res Methodol. 2019;19: 48. doi:10.1186/s12874-019-0688-x

27. Bouter L. Why research integrity matters and how it can be improved. Account Res. 2023;0: 1–10. doi:10.1080/08989621.2023.2189010

28. Gurwitz D, Milanesi E, Koenig T. Grant Application Review: The Case of Transparency. PLoS Biol. 2014;12: e1002010. doi:10.1371/journal.pbio.1002010

29. Horbach SPJM, Tijdink JK, Bouter L. Research funders should be more transparent: a plea for open applications. R Soc Open Sci. 2022;9: 220750. doi:10.1098/rsos.220750

30. Schweiger G. Can’t We Do Better? A cost-benefit analysis of proposal writing in a competitive funding environment. PloS One. 2023;18: e0282320. doi:10.1371/journal.pone.0282320

31. Ceci SJ, Kahn S, Williams WM. Exploring Gender Bias in Six Key Domains of Academic Science: An Adversarial Collaboration. Psychol Sci Public Interest. 2023;24: 15–73. doi:10.1177/15291006231163179

32. Schmaling KB, Gallo SA. Gender differences in peer reviewed grant applications, awards, and amounts: a systematic review and meta-analysis. Res Integr Peer Rev. 2023;8: 2. doi:10.1186/s41073-023-00127-3

33. Weber-Main AM, McGee R, Boman KE, Hemming J, Hall M, Unold T, et al. Grant application outcomes for biomedical researchers who participated in the National Research Mentoring Network’s Grant Writing Coaching Programs. PLOS ONE. 2020;15: e0241851. doi:10.1371/journal.pone.0241851

34. Guyer RA, Schwarze ML, Gosain A, Maggard-Gibbons M, Keswani SG, Goldstein AM. Top ten strategies to enhance grant-writing success. Surgery. 2021;170: 1727–1731. doi:10.1016/j.surg.2021.06.039

35. Bol T, de Vaan M, van de Rijt A. The Matthew effect in science funding. Proc Natl Acad Sci U S A. 2018;115: 4887–4890. doi:10.1073/pnas.1719557115

36. Heyard R, Ott M, Salanti G, Egger M. Rethinking the Funding Line at the Swiss National Science Foundation: Bayesian Ranking and Lottery. Stat Public Policy. 2022;9: 110–121. doi:10.1080/2330443X.2022.2086190

37. Qussini S, MacDonald RS, Shahbal S, Dierickx K. Blinding Models for Scientific Peer-Review of Biomedical Research Proposals: A Systematic Review. J Empir Res Hum Res Ethics. 2023;18: 250–262. doi:10.1177/15562646231191424

38. Grant J. The allocation of scientific grants should be a science. In: Times Higher Education (THE) [Internet]. 15 Jun 2017 [cited 30 Dec 2023]. Available: https://www.timeshighereducation.com/opinion/allocation-scientific-grants-should-be-science

39. Severin A, Egger M. Research on research funding: an imperative for science and society. Br J Sports Med. 2021;55: 648–649. doi:10.1136/bjsports-2020-103340

40. Azoulay P, Li D. Scientific Grant Funding. National Bureau of Economic Research; 2020. doi:10.3386/w26889

41. Bendiscioli S. The troubles with peer review for allocating research funding. EMBO Rep. 2019;20: e49472. doi:10.15252/embr.201949472

